# Adherence to Life’s Essential 8 is associated with delayed white matter aging

**DOI:** 10.1101/2024.12.02.24318274

**Authors:** Li Feng, Zhenyao Ye, Yezhi Pan, Rozalina G. McCoy, Braxton D. Mitchell, Peter Kochunov, Paul M. Thompson, Jie Chen, Menglu Liang, Thu T. Nguyen, Edmond Shenassa, Yan Li, Travis Canida, Hongjie Ke, Hwiyoung Lee, Song Liu, L. Elliot Hong, Chixiang Chen, David K.Y. Lei, Shuo Chen, Tianzhou Ma

## Abstract

**Importance:** The American Heart Association introduced Life’s Essential 8 (LE8) as a checklist of healthy lifestyle factors to help older individuals maintain and improve cardiovascular health and live longer. How LE8 can foster healthy brain aging and interact with genetic risk factors to render the aging brain less vulnerable to dementia is not well understood.

**Objective:** To investigate the impact of LE8 on the white matter brain aging and the moderating effects of the *APOE4* allele.

**Design, Setting, and Participants:** This cross-sectional study uses genetic, imaging, and other health-related data collected in the UK Biobank cohort. Participants included non-pregnant whites with LE8 variables, diffusion tensor imaging data, and genetic data on *APOE4* available, and excluded the extreme white matter hyperintensities. The baseline assessment was performed from 2006 to 2010. The diffusion tensor imaging data were collected since 2014.

**Exposures:** LE8 variables, encompassing diet, physical activity, smoking, sleep, body mass index, lipids, hemoglobin, and blood pressure.

**Main Outcomes and Measures:** The white matter brain age was predicted from regional fractional anisotropy measures derived from diffusion tensor imaging data using the random forest regression method. The outcome white matter brain age gap was calculated by subtracting individuals’ chronological age from their predicted brain age.

**Results:** The analysis included 9,430 women and 9,387 men (mean age 55.45 [SD: 7.46] years). Higher LE8 scores were associated with lower white matter brain age gap, indicating delayed brain aging. The findings are consistent for each of the individual LE8 variables. The effect was stronger among non-*APOE4* carriers (124 days younger per 10-point increase, 95% CI, 102 to 146 days; P<0.001) than *APOE4* carriers (84 days younger per 10-point increase, 95% CI, 47 to 120 days; P<0.001). Notably, early middle-aged women with *APOE4* showed significant interactions between LE8 scores and brain aging (P interaction = 0.048), not observed in men.

**Conclusions and Relevance:** Adherence to LE8 is associated with delayed brain aging, moderated by genetic factors such as *APOE4*. These findings highlight the potential of behavioral and lifestyle interventions in reducing dementia risk, emphasizing tailored prevention plans for those with different genetic predispositions to dementia and sex.

## Introduction

The American Heart Association introduced Life’s Essential 8 (LE8) as a comprehensive set of eight metrics that reflect health behaviors that support cardiovascular health (CVH)^1,2^, with the aim to help older individuals maintain CVH and live longer and healthier. These eight measures are categorized into two major areas: health behaviors (eating healthier foods, being more active, quitting tobacco, getting healthy sleep) and health factors (managing weight, controlling cholesterol, managing blood glucose, managing blood pressure). Beyond its association with CVH, LE8 is increasingly recognized for its impact on neurological health, including reduced risk of dementia (examined in association with LE8’s precursor, Life’s Simple 7 (LS7)^3^), potentially mediated by improved vascular health^4,5^. Recent studies linked higher LE8 scores with neuroimaging markers of better brain health (total brain volume, white matter hyperintensity, and hippocampal volume)^6^, better cognitive function^6–8^, and lower burden of cerebral small vessel disease^9^. As people age, the brain undergoes many changes including reduced neurogenesis, impaired synaptic connections, increased inflammation and among others^10^, which have been the major cause of cognitive decline in the elderly and the major risk factor of dementia. How LE8 can foster healthy brain aging and render the aging brain less vulnerable to dementia is not well known.

‘Brain age’ aims to predict chronological age from structural or functional neuroimaging features using a machine learning algorithm^11,12^. Brain Age Gap (BAG), which measures the discrepancy between chronological age and brain age^13,14^, has been linked to physical and brain health^13–18^, and serves as a precursor to cognitive decline and potential neurodegenerative diseases ^19^. Specifically, white matter (WM), the primary pathway for communication between different brain regions, is widely used to predict brain age as its structure and integrity significantly change with age. WM microstructural integrity has also been recognized as an early biomarker for the prodromal stage of Alzheimer’s disease (AD), dementia, and other neurodegenerative disorders^20–22^. Here, we focus on age-related WM microstructural changes and employ a machine learning algorithm^11,12,23^ to predict WM brain age from regional fractional anisotropy (FA) measures. Derived from diffusion tensor imaging (DTI) data, FA was shown as the primary factor contributing to cognitive aging rather than other neuroimaging markers^24^. In this study, we investigate how potential lifestyle-promoting factors like LE8 impact WM BAG.

In addition to environmental and lifestyle factors, multiple genetic variants^25,26^ have been implicated in brain aging and dementia risk^27^. The *APOE4* allele, among the strongest prevalent genetic risk factor of AD, is associated with an increased risk for AD by promoting the accumulation of beta-amyloid plaques and tau protein abnormalities, causing inflammation, compromising vascular health, and reducing neuroprotection in the brain^28–31^. However, the impact of *APOE4* on brain aging has been inconsistent across studies^6,32,33^. Some research also showed that *APOE4* may mitigate the protective effect from the lifestyle factors^34^, and sex may exhibit different effects^35^.

To fill the gap, here we conducted a study integrating genetic, neuroimaging, biomarker and questionnaire data from UK Biobank (UKB) to investigate the effect of LE8 (overall and of each lifestyle factor separately) on WM brain aging and examine whether these effects between LE8 and WM brain aging are modified by APOE4 status (a Gene x Environment interaction effect) in an age and sex-dependent manner (Figure 1). We hypothesized that the inconsistent associations between *APOE4* and brain aging are driven, in part, by its interaction with lifestyle factors such as the LE8. These data can provide mechanistic insights into how genetics and lifestyle factors jointly influence brain aging and cognitive impairment, ultimately informing personalized medicine plans for those at risk for AD and related dementia.

**Figure 1.**
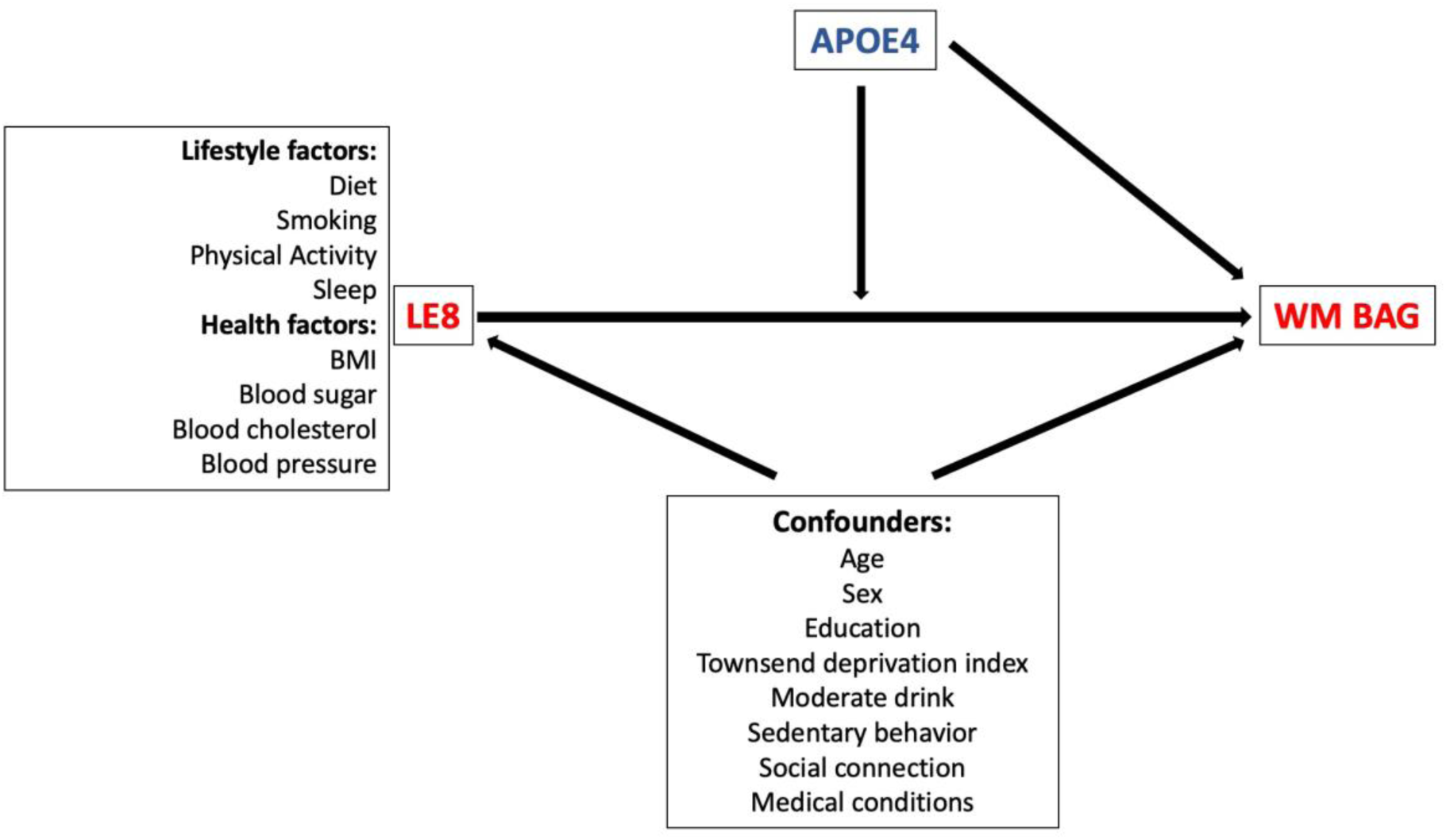
Directed Acyclic Graph (DAG): Illustrating the association between life’s essential 8 (LE8) and white matter brain age gap (WM BAG), controlling for confounders and modulated by the effect of APOE4 genotype.

## Methods

This cross-sectional study utilizes data from the UK Biobank (UKB), a population-based cohort study of more than 500,000 individuals aged 40 to 69 years in 22 recruitment centers across the UK, initially recruited from 2006 and 2010. The brain imaging phenotypic data collection began in 2014 for approximately 40,000 UKB participants^36^.

We focused specifically on non-pregnant white (predominantly European) participants to avoid bias of imbalanced data and reduce cross-population heterogeneity in training the brain age prediction model, consistent with our prior studies^23,37^. We calculated the LE8 score for those with complete data on all LE8 variables available (N=285,096). We estimated the outcome WM BAG for participants with available FA data and without extreme WM hyperintensities (N=30,375), to prevent distortions in structural brain measures^38^ (see Section: White matter brain age gap estimation). We investigated the main effect of LE8 on WM BAG among N=18,817 participants with both data available. We further considered participants with genetic data on APOE4 available to investigate the main effect of APOE4 on WM BAG (N=28,874) and the APOE4 x LE8 interaction effect (N=18,259). Details of the inclusion and exclusion criteria, data processing, and steps of the multiple statistical methods are shown in Figure S1.

### Neuroimaging data

The outcome, WM BAG, was computed based on the FA measures collected from DTI data from the UKB imaging assessment^23^. This study focuses on 39 WM FA tracts covering multiple brain regions (see the list of 39 regional WM FA measures in Table S1). Using the ENIGMA protocol, the average value for each white matter tract in the brain was computed using Tract-Based Spatial Statistics analysis applied to the DTI FA images^39,40^. Water diffusion is directionally restricted in healthy white matter, and FA values, which range from 0 (completely random diffusion, indicating damaged or less structured tissue) to 1 (highly directional diffusion, reflecting intact white matter), quantify the level of anisotropy in a diffusion process and the integrity of the white matter^41–43^.

### Assessment of Life’s Essential 8

The LE8 score is aligned with guidance from the American Heart Association^2^ and previous studies^1,44,45^ and comprises eight key metrics representing lifestyle behaviors (diet, physical activity, smoking, sleep) and health factors (body mass index (BMI), lipids (non-HDL cholesterol), hemoglobin A1c (HbA1c), and blood pressure (SBP and DBP). Diet quality scoring criteria are based on the Dietary Approaches to Stop Hypertension (DASH)-style diet. Each metric is assessed on a scale of 0 to 100, with the LE8 score calculated as the mean of these eight individual scores without weighted adjustments. The LE8 score was further categorized into three levels: scores ≥80 (high), scores ≥50 and <80 (middle), and scores <50 (low). Details on how we measure the LE8 metrics can be found in Tables S2 and S3.

### *APOE* Genotype

Individuals with *APOE* ε3/ε4 or ε4/ε4 genotypes were classified as *APOE4* carriers, while others (*APOE* ε2/ε2, ε2/ε3, ε3/ε3) were considered non-carriers. For analyzing the main effect of *APOE* on BAG, we further grouped *APOE* status into *APOE2* carriers (*APOE* ε2/ ε2, *APOE* ε2/ ε3), *APOE3* homozygotes (*APOE* ε3/ε3), and *APOE4* carriers (*APOE* ε3/ε4 or *APOE* ε4/ε4) (Details in supplementary section).

### Main Covariates

We included potential confounding factors from baseline in UKB: age, sex (females and males), education, Townsend Deprivation Index, household income, moderate alcohol consumption (women: ≤1 unit/day, men: ≤2 unit/day, yes/no), sedentary behavior, self-related social connection, and baseline health conditions (hypertension, cardiovascular diseases, type 2 diabetes, cancer, brain diseases) (Details in supplementary section).

### Statistical Analysis

We applied the random forest (RF) regression method to predict WM brain age and estimate WM BAG from the FA data. Following a similar data splitting scheme as our previous studies^13,14^, we first trained a RF regression model to generate a function to estimate unbiased brain age from regional FA measures in a training set of non-disease participants (those who were not previous or current smokers, and did not have hypertension, cardiovascular disease, diabetes, or brain diseases). The parameters of the RF regression were tuned based on the coefficients of determination (R) between the chronological age and estimated brain age and mean absolute error (MAE) criteria to achieve the optimal predictive performance using a 5-fold cross-validation. RF regression also selected a set of FA features that have the most significant impact on brain aging. The locked model was then applied to the testing samples including both disease and non-disease participants to predict their brain age. The WM BAG was calculated by subtracting individuals’ chronological age from their predicted brain age. Age-dependent bias has been found in brain age estimates across many clinical studies^14,46,47^ so we removed this brain age prediction bias from WM BAG and evaluated the performance of our correction method using the MAE.

We investigated the main effects of total LE8 score (0-100), individual LE8 components, categorized LE8 score (low, middle, high), as well as the main effect of *APOE* status, on WM BAG using general linear regression models. Model 1 adjusted for age and sex. Model 2 further adjusted for education, Townsend deprivation index, moderate alcohol consumption, sedentary behavior, and social connection. Model 3 further adjusted for the prevalent diseases. The inverse probability weighting (IPW) method^48^ was employed as an alternative method to mitigate the risk of confounding and provide potential causal evidence for the association between LE8 and WM BAG. We then investigated the potential interaction between lifestyle and *APOE4* to see how *APOE4* gene might alter the effect of LE8 on WM BAG. We also evaluated the effect of LE8 on WM BAG stratified by *APOE4* status. All these analyses were further stratified by baseline age (40-49, 50-59, 60-69) and sex (female or male).

All statistical analyses were conducted using R (version 4.0.5)^49^. R packages, including “*MICE*” (version 3.14.0), “*Hmisc*” (version 5.1.0) were used to perform multiple imputation by chained equations and inverse probability weighting analyses.

## Results

The overall sample used to examine the association between LE8 and the BAG included 18,817 white participants with a mean (SD) age of 55.45 (7.46) years of whom 49.22% were women. The mean (SD) LE8-total score (ranging from 0-100) was 71.90 (10.34). Participants in the middle and high LE8 group (≥80) were younger, more likely to be female, to have a college degree, greater household income, lower Townsend deprivation index, greater active social activity, and more likely to report “no or moderate” alcohol consumption compared to the low LE8 group (<50) (Table 1). In the Gene-Environment interaction sub-sample (N=18,259) exploring the effect modification of the *APOE4* allele, 25.82% were *APOE4* carriers (Table S5). *APOE4* carriers were younger, more often women, and with lower lipid levels but higher HbA1c and blood pressure levels (Table S5).

**Table 1.**
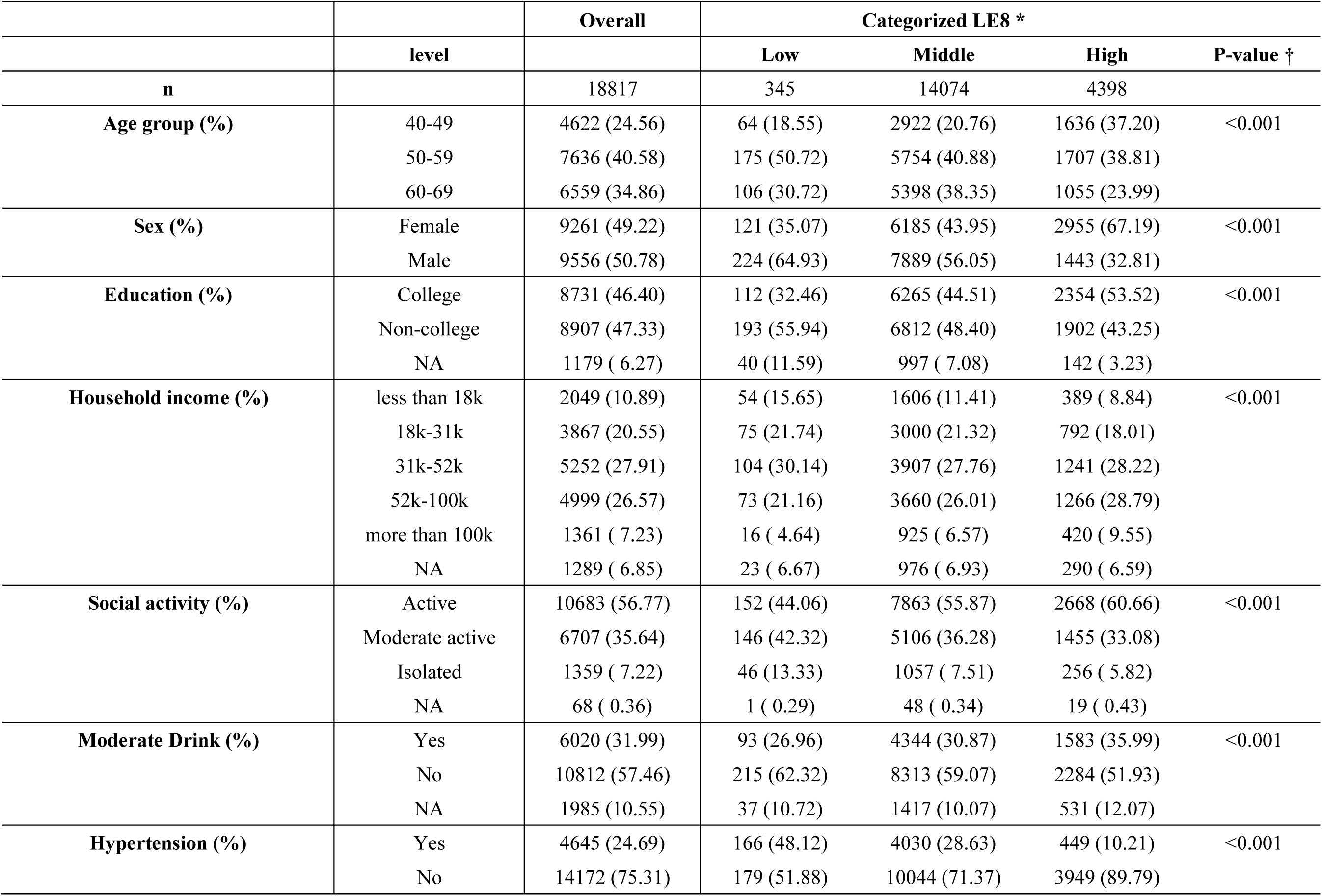
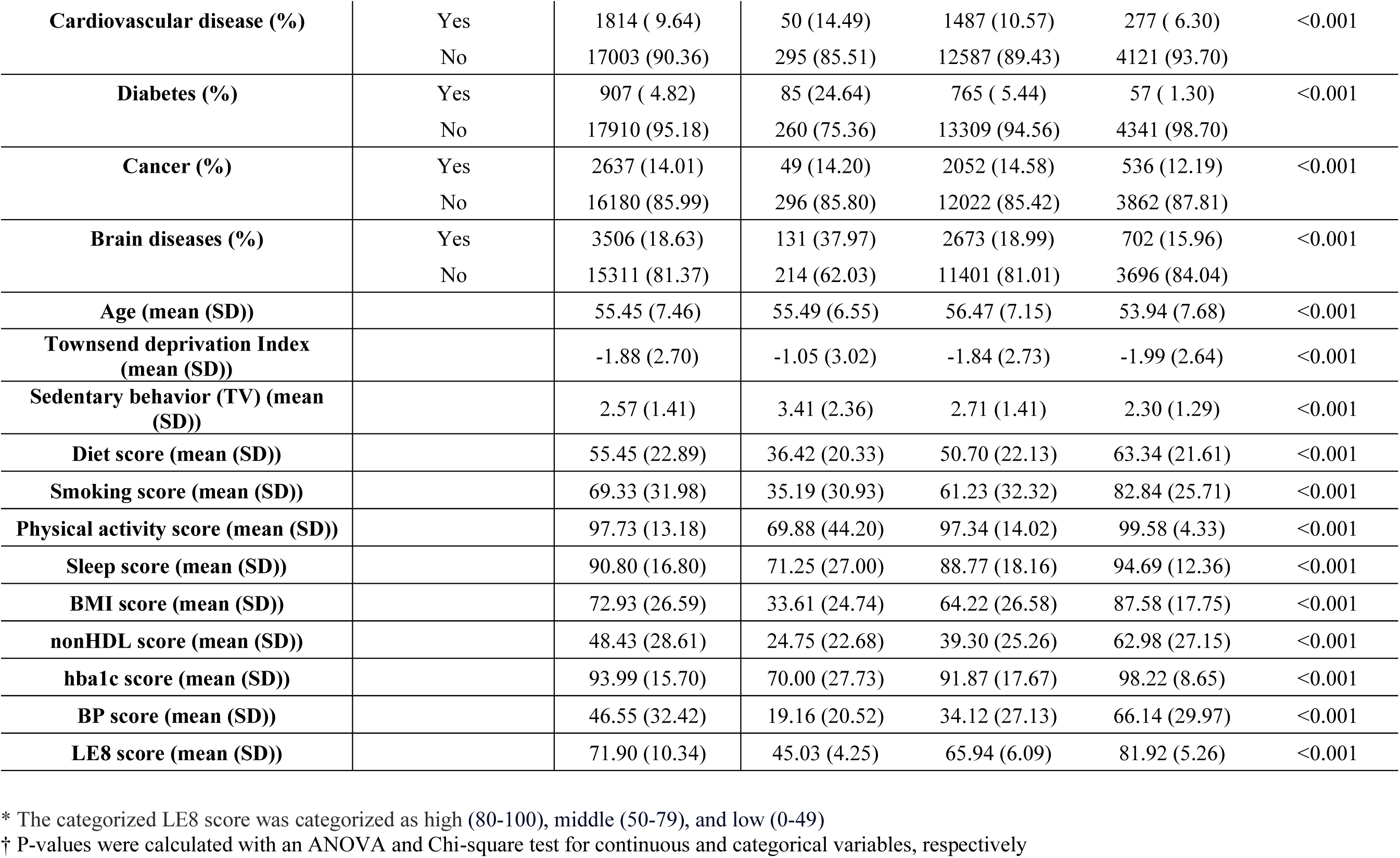
Baseline characteristics of participants in the UK Biobank by overall and categorized life’s essential 8 (LE8) (low, middle, and high) in analyzing the main effect of LE8 on the white matter (WM) brain age gap (BAG).

|  |  | Overall | Categorized LE8 * |  |  |  |
| --- | --- | --- | --- | --- | --- | --- |
|  | level |  | Low | Middle | High | P-value † |
| **n** |  | 18817 | 345 | 14074 | 4398 |  |
| **Age group (%)** | 40-49 | 4622 (24.56) | 64 (18.55) | 2922 (20.76) | 1636 (37.20) | <0.001 |
|  | 50-59 | 7636 (40.58) | 175 (50.72) | 5754 (40.88) | 1707 (38.81) |  |
|  | 60-69 | 6559 (34.86) | 106 (30.72) | 5398 (38.35) | 1055 (23.99) |  |
| **Sex (%)** | Female | 9261 (49.22) | 121 (35.07) | 6185 (43.95) | 2955 (67.19) | <0.001 |
|  | Male | 9556 (50.78) | 224 (64.93) | 7889 (56.05) | 1443 (32.81) |  |
| **Education (%)** | College | 8731 (46.40) | 112 (32.46) | 6265 (44.51) | 2354 (53.52) | <0.001 |
|  | Non-college | 8907 (47.33) | 193 (55.94) | 6812 (48.40) | 1902 (43.25) |  |
|  | NA | 1179 ( 6.27) | 40 (11.59) | 997 ( 7.08) | 142 ( 3.23) |  |
| **Household income (%)** | less than 18k | 2049 (10.89) | 54 (15.65) | 1606 (11.41) | 389 ( 8.84) | <0.001 |
|  | 18k-31k | 3867 (20.55) | 75 (21.74) | 3000 (21.32) | 792 (18.01) |  |
|  | 31k-52k | 5252 (27.91) | 104 (30.14) | 3907 (27.76) | 1241 (28.22) |  |
|  | 52k-100k | 4999 (26.57) | 73 (21.16) | 3660 (26.01) | 1266 (28.79) |  |
|  | more than 100k | 1361 ( 7.23) | 16 ( 4.64) | 925 ( 6.57) | 420 ( 9.55) |  |
|  | NA | 1289 ( 6.85) | 23 ( 6.67) | 976 ( 6.93) | 290 ( 6.59) |  |
| **Social activity (%)** | Active | 10683 (56.77) | 152 (44.06) | 7863 (55.87) | 2668 (60.66) | <0.001 |
|  | Moderate active | 6707 (35.64) | 146 (42.32) | 5106 (36.28) | 1455 (33.08) |  |
|  | Isolated | 1359 ( 7.22) | 46 (13.33) | 1057 ( 7.51) | 256 ( 5.82) |  |
|  | NA | 68 ( 0.36) | 1 ( 0.29) | 48 ( 0.34) | 19 ( 0.43) |  |
| **Moderate Drink (%)** | Yes | 6020 (31.99) | 93 (26.96) | 4344 (30.87) | 1583 (35.99) | <0.001 |
|  | No | 10812 (57.46) | 215 (62.32) | 8313 (59.07) | 2284 (51.93) |  |
|  | NA | 1985 (10.55) | 37 (10.72) | 1417 (10.07) | 531 (12.07) |  |
| **Hypertension (%)** | Yes | 4645 (24.69) | 166 (48.12) | 4030 (28.63) | 449 (10.21) | <0.001 |
|  | No | 14172 (75.31) | 179 (51.88) | 10044 (71.37) | 3949 (89.79) |  |
| **Cardiovascular disease (%)** | Yes | 1814 ( 9.64) | 50 (14.49) | 1487 (10.57) | 277 ( 6.30) | <0.001 |
|  | No | 17003 (90.36) | 295 (85.51) | 12587 (89.43) | 4121 (93.70) |  |
| **Diabetes (%)** | Yes | 907 ( 4.82) | 85 (24.64) | 765 ( 5.44) | 57 ( 1.30) | <0.001 |
|  | No | 17910 (95.18) | 260 (75.36) | 13309 (94.56) | 4341 (98.70) |  |
| **Cancer (%)** | Yes | 2637 (14.01) | 49 (14.20) | 2052 (14.58) | 536 (12.19) | <0.001 |
|  | No | 16180 (85.99) | 296 (85.80) | 12022 (85.42) | 3862 (87.81) |  |
| **Brain diseases (%)** | Yes | 3506 (18.63) | 131 (37.97) | 2673 (18.99) | 702 (15.96) | <0.001 |
|  | No | 15311 (81.37) | 214 (62.03) | 11401 (81.01) | 3696 (84.04) |  |
| **Age (mean (SD))** |  | 55.45 (7.46) | 55.49 (6.55) | 56.47 (7.15) | 53.94 (7.68) | <0.001 |
| **Townsend deprivation Index (mean (SD))** |  | -1.88 (2.70) | -1.05 (3.02) | -1.84 (2.73) | -1.99 (2.64) | <0.001 |
| **Sedentary behavior (TV) (mean (SD))** |  | 2.57 (1.41) | 3.41 (2.36) | 2.71 (1.41) | 2.30 (1.29) | <0.001 |
| **Diet score (mean (SD))** |  | 55.45 (22.89) | 36.42 (20.33) | 50.70 (22.13) | 63.34 (21.61) | <0.001 |
| **Smoking score (mean (SD))** |  | 69.33 (31.98) | 35.19 (30.93) | 61.23 (32.32) | 82.84 (25.71) | <0.001 |
| **Physical activity score (mean (SD))** |  | 97.73 (13.18) | 69.88 (44.20) | 97.34 (14.02) | 99.58 (4.33) | <0.001 |
| **Sleep score (mean (SD))** |  | 90.80 (16.80) | 71.25 (27.00) | 88.77 (18.16) | 94.69 (12.36) | <0.001 |
| **BMI score (mean (SD))** |  | 72.93 (26.59) | 33.61 (24.74) | 64.22 (26.58) | 87.58 (17.75) | <0.001 |
| **nonHDL score (mean (SD))** |  | 48.43 (28.61) | 24.75 (22.68) | 39.30 (25.26) | 62.98 (27.15) | <0.001 |
| **hba1c score (mean (SD))** |  | 93.99 (15.70) | 70.00 (27.73) | 91.87 (17.67) | 98.22 (8.65) | <0.001 |
| **BP score (mean (SD))** |  | 46.55 (32.42) | 19.16 (20.52) | 34.12 (27.13) | 66.14 (29.97) | <0.001 |
| **LE8 score (mean (SD))** |  | 71.90 (10.34) | 45.03 (4.25) | 65.94 (6.09) | 81.92 (5.26) | <0.001 |

\* The categorized LE8 score was categorized as high (80-100), middle (50-79), and low (0-49)
† P-values were calculated with an ANOVA and Chi-square test for continuous and categorical variables, respectively

### Estimation of the outcome WM BAG

The optimal random forest regression model selected 16 FA measures for BAG estimation (Table S1). After correcting for age bias on the white matter BAG, the adjusted predicted BAG achieved R= 0.89, MAE = 2.74 years in both the disease test dataset and the non-disease test dataset (Figure S2). The disease group was, on average, 0.28 years (95%CI= 0.19 to 0.36; *p* = 4.91E-10) older in WM brain age than the non-disease group.

### Main effect of LE8 on WM BAG

The regression analyses revealed significant associations between LE8 and WM BAG for both continuous and categorized LE8 (Table 2). Higher LE8 score was associated with a lower WM BAG (β_model1_ = −0.42, β_model2_ = −0.39, β_model3_ = −0.31, all *p*<0.001), indicating delayed brain aging (153, 142 and 113 days younger brain age per 10-point increase for model 1, 2 and 3, respectively). The middle LE8 group (β = −0.88 (−321.20 days), 95% CI: −1.23 to −0.53 (−448.95 to −193.45 days), *p*<0.001) and the high LE8 group (β= −1.34 (−489.10 days), 95% CI: −1.71 to −0.99 (−624.15 to −361.35 days), *p*<0.001) showed a significantly lower WM BAG compared to the low LE8 group (as reference) in model 3. This pattern was held across all models, adjusting for demographics, socioeconomic status, and medical conditions. IPW analysis validated the effect of LE8 groups on WM BAG from our main analysis and provided potentially causal evidence (Table S6). Consistent beneficial associations in all models were also observed for individual LE8 factors including a favorable diet, no smoking, higher level of physical activity, favorable BMI, and normal blood pressure and HbA1c levels (Table S7 and Figure 2).

**Figure 2.**
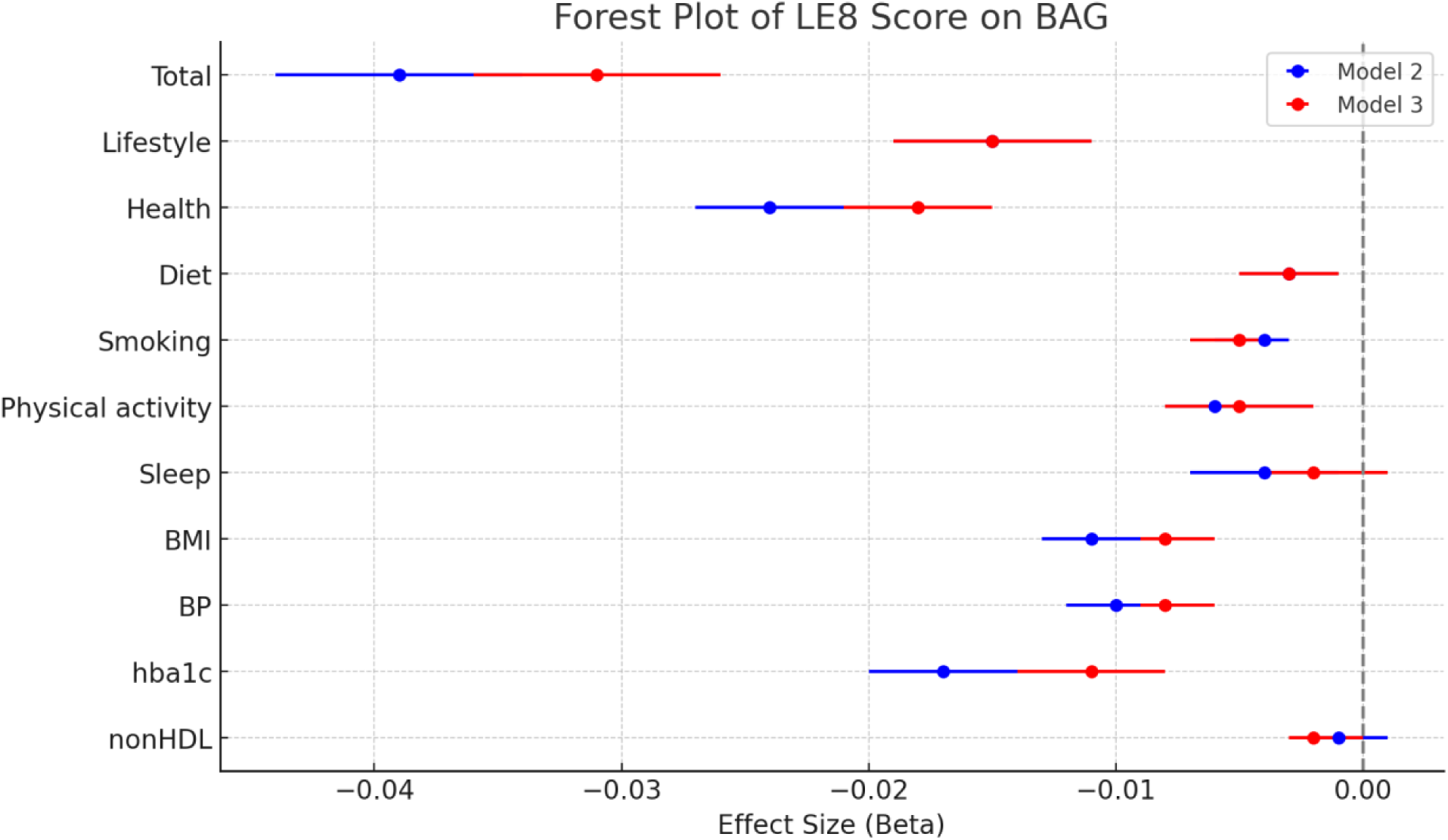
Forest plot of the effect sizes with 95% confidence intervals for joint and individual life’s essential 8 (LE8) scores on white matter brain age gap (BAG) in models 2 and 3. Model 2: model 1+ education, Townsend deprivation index, moderate drink, sedentary behavior, social connection. Model 3: model 2+ medical conditions (hypertension, cardiovascular disease, cancer, brain diseases). BMI: body mass index. BP: blood pressure.

**Table 2.**
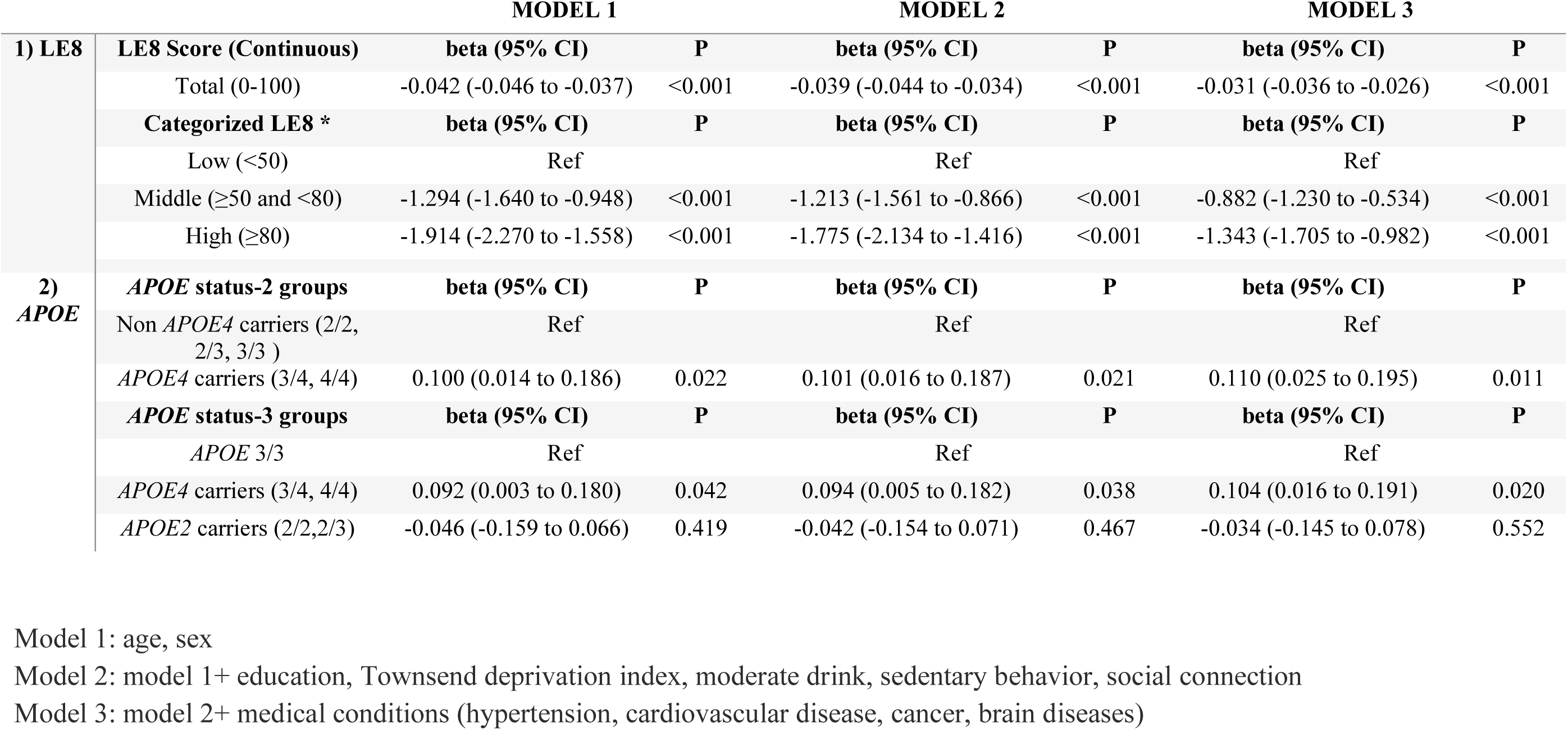
Main effects of 1) joint life’s essential 8 (LE8) and 2) APOE status on white matter Brain Age Gap by using the general linear regression model. 95% CI: 95% confidence interval.

The steepest decline in WM BAG with increasing LE8 score was observed in the 50-59 age group (Figure S3A, *p* for interaction (40-49 vs. 50-59) <0.001). There was no sex difference in the association between LE8-total score and WM BAG in each age group (all *p* _interaction_>0.05) (Figure S3B).

### Main and modification effect of *APOE4* genotype on WM BAG

*APOE4* carriers exhibited a significantly greater WM BAG compared to non-*APOE4* carriers across all three models (β_model1_ = 0.10, β_model2_ = 0.10, β_model3_ = 0.11, *p*=0.02, 0.02 and 0.01), indicating accelerated brain aging. In contrast, *APOE3/3* and *APOE2* carriers (ε2/ε2, ε2/ε3) did not show any significant difference in WM BAG (Table 2). Therefore, for subsequent analyses, we only focused on the comparison between *APOE4* carriers and non-carriers. The modification effects of the *APOE4* genotype revealed a complex pattern. No overall interaction between LE8 and *APOE4* status was observed; however, a few single LE8 x *APOE4* interactions showed marginal significance among women and men (Table S8). Specifically, we observed a significant interaction in women aged 40-49 (*p* _interaction_ = 0.048) (Figure 3). Stratified analyses by *APOE4* status further revealed that non-*APOE4* carriers consistently benefit from higher LE8 scores with reduced WM BAG across all age groups, whereas *APOE4* carriers exhibit variable and generally weaker responses (Table S9 & S10). The overall observed patterns indicate that health behaviors/lifestyle factors may differentially affect brain aging and this impact is further modulated by *APOE4* status across sexes.

**Figure 3.**
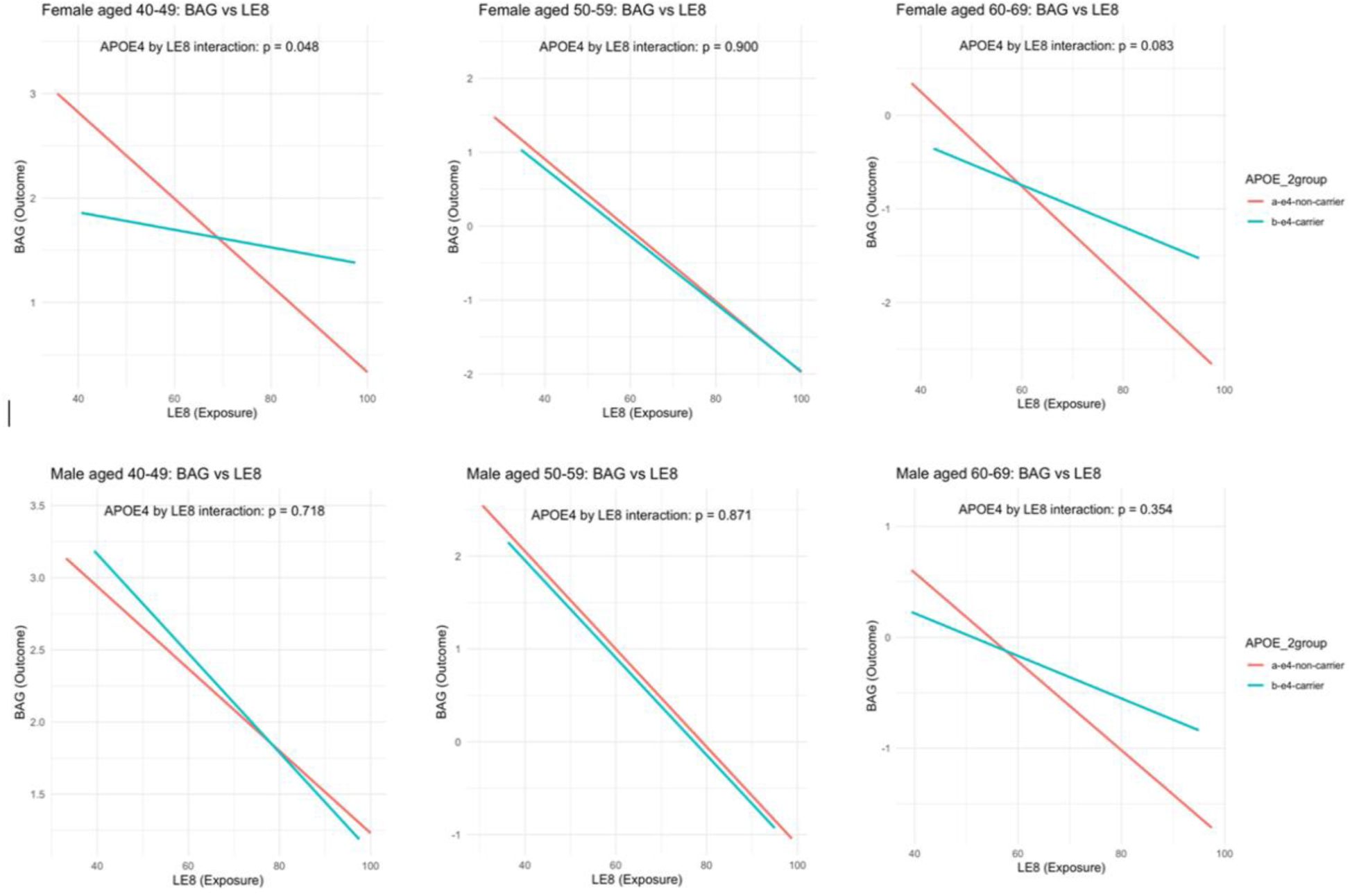
Interaction effects of *APOE4* and life’s essential 8 (LE8) scores on white matter (WM) brain age gap (BAG) across different age and sex groups.

### Risk ratios of dementia among *APOE4* and LE8 combined groups

As white matter brain aging is an early biomarker for dementia, we also calculated the crude and age-adjusted risk factors for all-cause dementia in different groups classified by *APOE4* status and LE8 score in UKB (Table S4). Overall, *APOE4* carriers had a higher risk of all-cause dementia in women and men compared to non-*APOE4* carriers. *APOE4* carriers with high LE8 (>=70) in women (crude RR: 0.62, *p*<0.001) and men (crude RR: 0.82, *p*=0.003) showed a significant reduction in all-cause dementia compared with those with low LE8 (<70), though this association lost significance after age adjustment (Figure S4 and Table S11).

## Discussion

Our study, conducted using rich genetic, imaging, and other health-related data from the UKB, advances the understanding of how LE8 is associated with white matter brain aging and how this effect is modified by *APOE4* status in a large prospective cohort in the UK. Our findings highlight that higher LE8 scores were associated with delayed white matter brain aging. This was further influenced by the *APOE4* allele’s modification effects, where non-*APOE4* carriers showed a consistent delayed brain aging across all levels of LE8, and *APOE4* carriers displayed diminished effects. Notably, our findings highlighted sex and age disparities, particularly among *APOE4* carriers, where the expected protective influence of LE8 was not uniformly evident.

Our findings linking LE8 to delayed white matter brain aging have not been previously described and were built on prior studies that found associations of healthy behaviors/lifestyles (LS7, LE8, and related) with better cognition, lower dementia risk, and better micro-/macro-brain structures^6,9,50–56^. Collectively, we highlight the importance of adherence to a healthy lifestyle and behavior in maintaining brain integrity across various demographics and ages. Our findings also underscore the complexity of the gene-environment interaction in neurodegenerative risk, especially in the context of the *APOE4* allele. While a higher LE8 score is associated with a lower WM BAG regardless of *APOE4* status, we observed an attenuated effect size of LE8 in *APOE4* carriers. This attenuation suggests a potential limit to the neuroprotective effects of the lifestyle factors in individuals carrying the *APOE4* allele. This observation aligns with prior studies examining white matter integrity^57^ and dementia^6,58–60^. We have previously highlighted the moderating effect of *APOE4* on the relationship between plasma metabolites and white matter microstructural integrity^61^, where low-density lipoprotein detrimentally affects white matter integrity in critical neural tracts among *APOE4* carriers, while they have a protective effect on white matter integrity among non-carriers. A sex-*APOE4* interaction exists in brain health^62,63^. In this study, we observed that women and men experienced varying degrees of brain health protection from positive LE8 score changes with or without the *APOE4* allele. Together, these findings highlight the need to incorporate healthy behaviors and lifestyles into strategies designed to slow down brain aging while also considering genetic predispositions and sex differences to optimize the effectiveness of such interventions.

Our study has several strengths. Firstly, it leverages large-scale imaging, genetic and biomarker data from the UK Biobank. Second, using machine learning to compute biological brain age introduces an innovative and potentially more accurate method of assessing brain aging, allowing us to detect subtle changes in brain aging that traditional methods might overlook. Additionally, we offer comprehensive LE8 metrics, including a wide range of modifiable behaviors and health factors, as a holistic approach. Moreover, the inclusion of the *APOE4* modification effect together with age- and sex-stratified analysis addresses an important aspect of personalized and precision medicine. Lastly, our approach is strengthened by using robust and rigorous statistical tools, including the IPW method.

This study also has some limitations. First, it includes potential selection bias due to the relatively healthy volunteers in the UK Biobank population^64^. Second, reliance on self-reported data collected at baseline for lifestyle factors could introduce recall or misclassification bias. Furthermore, unmeasured variables such as environmental factors and psychosocial stressors might influence outcomes. Finally, the limitation of tracking the changes of LE8 and brain aging over time may hamper our capacity to establish causal relationships. Future studies involving ongoing longitudinal cohorts, such as the Adolescent Brain Cognitive Development study^65^ and the Baltimore Longitudinal Study of Aging^66^, could provide more comprehensive insights.

## Conclusion

Our research provides evidence that Life’s Essential 8, is vital in slowing the progression of white matter brain aging, particularly in the context of the *APOE4* genotype and different sexes among 40- to 69-year-olds of European ancestry, in the UK Biobank. The implications of our findings are profound, suggesting that concerted efforts to healthy lifestyles and behaviors may significantly contribute to preserving brain health and delaying the aging process of the brain, with tailored prevention plans for those with different genetic predispositions to AD and related dementia and sex.

## Author Contributions

LF took the lead in performing the analysis and wrote the manuscript. ZY and YP processed the genetic and imaging data. LF, TM and SC conceptualized the idea. TM and SC supervised the project and took the lead in editing the manuscript. TM acquired the funding. RGM, BDM, PK, PMT, JC, ML, TN, ES, YL, TC, HK, HL, SL, EH, CC and DL contributed to manuscript writing and polishing. All authors provided critical feedback and helped to shape the research, analysis, and manuscript.

## Source of Funding

Research reported in this publication was supported by the National Institute on Drug Abuse (NIDA) of National Institute of Health under the award number 1DP1DA048968-01 to SC and TM, by the National Institute on Drug Abuse (NIDA) under the award number 1K01DA059603-01A1 to TM, by the University of Maryland MPower Brain Health and Human Performance seed grant to TM and PK, by the Grand Challenge Grant to TM, and EPIB Department Pilot Award to TM, TN and YL. RGM, SC, CC, JC, are TN are investigators at the University of Maryland-Institute for Health Computing, which is supported by funding from Montgomery County, Maryland and The University of Maryland Strategic Partnership: MPowering the State, a formal collaboration between the University of Maryland, College Park and the University of Maryland, Baltimore.

## Disclosures

In the last 36 months, RGM has received support from the National Institute of Diabetes and Digestive and Kidney Diseases (NIDDK) of the National Institutes of Health (NIH), National Institute on Aging (NIA) of the NIH, Patient Centered Outcomes Research Institute (PCORI), National Center for Advancing Translational Sciences (NCATS), and the American Diabetes Association. She serves as a consultant to EmmiEducate^®^ (Wolters Kluwer) and the Yale-New Haven Health System’s Center for Outcomes Research and Evaluation, and has received speaking honoraria and travel support from the American Diabetes Association.

## Data Availability Statement

The raw genetic and phenotypic data used for this study can be found in the UK Biobank (http://www.ukbiobank.ac.uk/)

## Data Availability

All data produced in the present study are available upon reasonable request to the authors.

## Reference

1. Hayman LL, Martyn-Nemeth P. A New Metric for Promoting Cardiovascular Health: Life’s Essential 8. J Cardiovasc Nurs. 2022;37(6):509–511.

2. Lloyd-Jones DM, Allen NB, Anderson CAM, et al. Life’s Essential 8: Updating and Enhancing the American Heart Association’s Construct of Cardiovascular Health: A Presidential Advisory From the American Heart Association. Circulation. 2022;146(5):e18–e43.

3. Hasbani NR, Ligthart S, Brown MR, et al. American Heart Association’s Life’s Simple 7: Lifestyle Recommendations, Polygenic Risk, and Lifetime Risk of Coronary Heart Disease. Circulation. 2022;145(11):808–818.

4. Wang R, Fratiglioni L, Kalpouzos G, et al. Mixed brain lesions mediate the association between cardiovascular risk burden and cognitive decline in old age: A population-based study. Alzheimers Dement. 2017;13(3):247–256.

5. Cox SR, Lyall DM, Ritchie SJ, et al. Associations between vascular risk factors and brain MRI indices in UK Biobank. Eur Heart J. 2019;40(28):2290–2300.

6. Zhou R, Chen HW, Li FR, Zhong Q, Huang YN, Wu XB. “Life’s Essential 8” Cardiovascular Health and Dementia Risk, Cognition, and Neuroimaging Markers of Brain Health. J Am Med Dir Assoc. 2023;24(11):1791–1797.

7. Liang K, Zhang X. Association between Life’s Essential 8 and cognitive function: insights from NHANES 2011-2014. Front Aging Neurosci. 2024;16:1386498.

8. Ferreira NV, Gonçalves NG, Szlejf C, et al. Optimal cardiovascular health is associated with slower cognitive decline. European Journal of Neurology. 2024;31(2):e16139.

9. Liu D, Cai X, Yang Y, et al. Association between Life’s Essential 8 and Cerebral Small Vessel Disease. Stroke Vasc Neurol. 2023.

10. Mattson MP, Arumugam TV. Hallmarks of Brain Aging: Adaptive and Pathological Modification by Metabolic States. Cell Metab. 2018;27(6):1176–1199.

11. Cole JH, Franke K. Predicting Age Using Neuroimaging: Innovative Brain Ageing Biomarkers. Trends Neurosci. 2017;40(12):681–690.

12. Cole JH, Marioni RE, Harris SE, Deary IJ. Brain age and other bodily ‘ages’: implications for neuropsychiatry. Mol Psychiatry. 2019;24(2):266–281.

13. Feng L, Ye Z, Mo C, et al. Elevated blood pressure accelerates white matter brain aging among late middle-aged women: a Mendelian Randomization study in the UK Biobank. J Hypertens. 2023;41(11):1811–1820.

14. Mo C, Wang J, Ye Z, et al. Evaluating the causal effect of tobacco smoking on white matter brain aging: a two-sample Mendelian randomization analysis in UK Biobank. Addiction. 2022.

15. Hogan AM, Vargha-Khadem F, Saunders DE, Kirkham FJ, Baldeweg T. Impact of frontal white matter lesions on performance monitoring: ERP evidence for cortical disconnection. Brain. 2006;129(8):2177–2188.

16. Gaser C, Franke K, Klöppel S, Koutsouleris N, Sauer H, Initiative AsDN. BrainAGE in mild cognitive impaired patients: predicting the conversion to Alzheimer’s disease. PloS one. 2013;8(6):e67346.

17. Kolbeinsson A, Filippi S, Panagakis Y, et al. Accelerated MRI-predicted brain ageing and its associations with cardiometabolic and brain disorders. Scientific Reports. 2020;10(1):1–9.

18. Ye Z, Mo C, Liu S, et al. Deciphering the causal relationship between blood pressure and regional white matter integrity: A two-sample Mendelian randomization study. J Neurosci Res. 2023;101(9):1471–1483.

19. Coelho A, Fernandes HM, Magalhaes R, et al. Signatures of white-matter microstructure degradation during aging and its association with cognitive status. Sci Rep. 2021;11(1):4517.

20. Kantarci K, Schwarz CG, Reid RI, et al. White matter integrity determined with diffusion tensor imaging in older adults without dementia: influence of amyloid load and neurodegeneration. JAMA Neurol. 2014;71(12):1547–1554.

21. Gold BT, Johnson NF, Powell DK, Smith CD. White matter integrity and vulnerability to Alzheimer’s disease: preliminary findings and future directions. Biochim Biophys Acta. 2012;1822(3):416–422.

22. Xiao D, Wang K, Theriault L, Charbel E, Alzheimer’s Disease Neuroimaging I. White matter integrity and key structures affected in Alzheimer’s disease characterized by diffusion tensor imaging. Eur J Neurosci. 2022;56(8):5319–5331.

23. Feng L, Ye Z, Mo C, et al. Elevated blood pressure accelerates white matter brain aging among late middle-aged women: a Mendelian Randomization study in the UK Biobank. J Hypertens. 2023.

24. Lee H, Chen C, Kochunov P, Hong LE, Chen S. A new multiple-mediator model maximally uncovering the mediation pathway: Evaluating the role of neuroimaging measures in age-related cognitive decline. The Annals of Applied Statistics. 2024;18(4):2775–2795, 2721.

25. Seshadri S, DeStefano AL, Au R, et al. Genetic correlates of brain aging on MRI and cognitive test measures: a genome-wide association and linkage analysis in the Framingham Study. BMC Med Genet. 2007;8 Suppl 1(Suppl 1):S15.

26. Brouwer RM, Klein M, Grasby KL, et al. Genetic variants associated with longitudinal changes in brain structure across the lifespan. Nat Neurosci. 2022;25(4):421–432.

27. Jansen IE, Savage JE, Watanabe K, et al. Genome-wide meta-analysis identifies new loci and functional pathways influencing Alzheimer’s disease risk. Nat Genet. 2019;51(3):404–413.

28. Lefterov I, Fitz NF, Lu Y, Koldamova R. APOEepsilon4 and risk of Alzheimer’s disease - time to move forward. Front Neurosci. 2023;17:1195724.

29. Palmer JM, Huentelman M, Ryan L. More than just risk for Alzheimer’s disease: APOE epsilon4’s impact on the aging brain. Trends Neurosci. 2023;46(9):750–763.

30. Scuteri A, Najjar SS, Muller D, et al. apoE4 allele and the natural history of cardiovascular risk factors. Am J Physiol Endocrinol Metab. 2005;289(2):E322–327.

31. Tini G, Scagliola R, Monacelli F, et al. Alzheimer’s Disease and Cardiovascular Disease: A Particular Association. Cardiol Res Pract. 2020;2020:2617970.

32. Lowe LC, Gaser C, Franke K, Alzheimer’s Disease Neuroimaging I. The Effect of the APOE Genotype on Individual BrainAGE in Normal Aging, Mild Cognitive Impairment, and Alzheimer’s Disease. PLoS One. 2016;11(7):e0157514.

33. Scheller E, Schumacher LV, Peter J, et al. Brain Aging and APOE epsilon4 Interact to Reveal Potential Neuronal Compensation in Healthy Older Adults. Front Aging Neurosci. 2018;10:74.

34. Kivipelto M, Rovio S, Ngandu T, et al. Apolipoprotein E epsilon4 magnifies lifestyle risks for dementia: a population-based study. J Cell Mol Med. 2008;12(6B):2762–2771.

35. Eid A, Mhatre I, Richardson JR. Gene-environment interactions in Alzheimer’s disease: A potential path to precision medicine. Pharmacol Ther. 2019;199:173–187.

36. Sudlow C, Gallacher J, Allen N, et al. UK biobank: an open access resource for identifying the causes of a wide range of complex diseases of middle and old age. PLoS Med. 2015;12(3):e1001779.

37. Ye Z, Mo C, Liu S, et al. Deciphering the causal relationship between blood pressure and regional white matter integrity: A two-sample Mendelian randomization study. J Neurosci Res. 2023.

38. Maniega SM, Valdes Hernandez MC, Clayden JD, et al. White matter hyperintensities and normal-appearing white matter integrity in the aging brain. Neurobiol Aging. 2015;36(2):909–918.

39. Smith SM, Jenkinson M, Johansen-Berg H, et al. Tract-based spatial statistics: voxelwise analysis of multi-subject diffusion data. Neuroimage. 2006;31(4):1487–1505.

40. Timpe JC, Rowe KC, Matsui J, Magnotta VA, Denburg NL. White matter integrity, as measured by diffusion tensor imaging, distinguishes between impaired and unimpaired older adult decision-makers: A preliminary investigation. J Cogn Psychol (Hove). 2011;23(6):760–767.

41. Voineskos AN, Rajji TK, Lobaugh NJ, et al. Age-related decline in white matter tract integrity and cognitive performance: a DTI tractography and structural equation modeling study. Neurobiology of aging. 2012;33(1):21–34.

42. Assaf Y, Pasternak O. Diffusion tensor imaging (DTI)-based white matter mapping in brain research: a review. J Mol Neurosci. 2008;34(1):51–61.

43. Garin-Muga A, Borro D. Review and Challenges of Brain Analysis through DTI Measurements. Stud Health Technol Inform. 2014;207:27–36.

44. Petermann-Rocha F, Deo S, Celis-Morales C, et al. An Opportunity for Prevention: Associations Between the Life’s Essential 8 Score and Cardiovascular Incidence Using Prospective Data from UK Biobank. Curr Probl Cardiol. 2023;48(4):101540.

45. Sun J, Li Y, Zhao M, et al. Association of the American Heart Association’s new “Life’s Essential 8” with all-cause and cardiovascular disease-specific mortality: prospective cohort study. BMC Med. 2023;21(1):116.

46. Le TT, Kuplicki RT, McKinney BA, et al. A Nonlinear Simulation Framework Supports Adjusting for Age When Analyzing BrainAGE. Front Aging Neurosci. 2018;10:317.

47. Beheshti I, Nugent S, Potvin O, Duchesne S. Bias-adjustment in neuroimaging-based brain age frameworks: A robust scheme. Neuroimage Clin. 2019;24:102063.

48. Shiba K, Kawahara T. Using Propensity Scores for Causal Inference: Pitfalls and Tips. J Epidemiol. 2021;31(8):457–463.

49. R.C. T. R: A language and environment for statistical computing. 2013.

50. Gardener H, Caunca M, Dong C, et al. Ideal Cardiovascular Health and Biomarkers of Subclinical Brain Aging: The Northern Manhattan Study. J Am Heart Assoc. 2018;7(16):e009544.

51. Fuhrmann D, Nesbitt D, Shafto M, et al. Strong and specific associations between cardiovascular risk factors and white matter micro- and macrostructure in healthy aging. Neurobiol Aging. 2019;74:46–55.

52. Austin TR, Nasrallah IM, Erus G, et al. Association of Brain Volumes and White Matter Injury With Race, Ethnicity, and Cardiovascular Risk Factors: The Multi-Ethnic Study of Atherosclerosis. J Am Heart Assoc. 2022;11(7):e023159.

53. Liu D, Cai X, Yang Y, et al. Associations of Life’s Simple 7 With Cerebral Small Vessel Disease. Stroke. 2022;53(9):2859–2867.

54. Keller JA, Kant IMJ, Slooter AJC, et al. Different cardiovascular risk factors are related to distinct white matter hyperintensity MRI phenotypes in older adults. Neuroimage Clin. 2022;35:103131.

55. Fu Y, Sun Y, Wang ZB, et al. Associations of Life’s Simple 7 with cerebral white matter hyperintensities and microstructural integrity: UK Biobank cohort study. Eur J Neurol. 2023;30(5):1200–1208.

56. Koohi F, Harshfield EL, Markus HS. Contribution of Conventional Cardiovascular Risk Factors to Brain White Matter Hyperintensities. J Am Heart Assoc. 2023;12(14):e030676.

57. Smith JC, Lancaster MA, Nielson KA, et al. Interactive effects of physical activity and APOE-epsilon4 on white matter tract diffusivity in healthy elders. Neuroimage. 2016;131:102–112.

58. Pa J, Aslanyan V, Casaletto KB, et al. Effects of Sex, APOE4, and Lifestyle Activities on Cognitive Reserve in Older Adults. Neurology. 2022;99(8):e789–e798.

59. Lourida I, Hannon E, Littlejohns TJ, et al. Association of Lifestyle and Genetic Risk With Incidence of Dementia. JAMA. 2019;322(5):430–437.

60. Guo J, Brickman AM, Manly JJ, et al. Association of Life’s Simple 7 with incident dementia and its modification by the apolipoprotein E genotype. Alzheimers Dement. 2021;17(12):1905–1913.

61. Ye Z, Pan Y, McCoy RG, et al. Contrasting association pattern of plasma low-density lipoprotein with white matter integrity in APOE4 carriers versus non-carriers. Neurobiol Aging. 2024;143:41–52.

62. Srisaikaew P, Chad JA, Mahakkanukrauh P, Anderson ND, Chen JJ. Effect of sex on the APOE4-aging interaction in the white matter microstructure of cognitively normal older adults using diffusion-tensor MRI with orthogonal-tensor decomposition (DT-DOME). Front Neurosci. 2023;17:1049609.

63. Hsu M, Dedhia M, Crusio WE, Delprato A. Sex differences in gene expression patterns associated with the APOE4 allele. F1000Res. 2019;8:387.

64. Fry A, Littlejohns TJ, Sudlow C, et al. Comparison of Sociodemographic and Health-Related Characteristics of UK Biobank Participants With Those of the General Population. Am J Epidemiol. 2017;186(9):1026–1034.

65. Casey BJ, Cannonier T, Conley MI, et al. The Adolescent Brain Cognitive Development (ABCD) study: Imaging acquisition across 21 sites. Dev Cogn Neurosci. 2018;32:43–54.

66. Ferrucci L. The Baltimore Longitudinal Study of Aging (BLSA): a 50-year-long journey and plans for the future. J Gerontol A Biol Sci Med Sci. 2008;63(12):1416–1419.

