## Supplementary methods for "Adherence to Life’s Essential 8 is associated with delayed white matter aging"

### *APOE* Genotype

The genotype data were assayed with UK BiLEVE Axiom Array and with UK Biobank Axiom™ platforms[1,2]. Our secondary analysis focused on white individuals without kinship or more than 2% missing genotypes processed using PLINK[3] (version 1.9, www.cog-genomics.org/plink/1.9/). The *APOE* genotype of participants was determined by the two *APOE* isoform coding single nucleotide polymorphisms[4], rs429358 and rs7412, located on chromosome 19. Individuals were categorized as *APOE4*-carriers if they possessed the *APOE* ε3/ε4 or *APOE* ε4/ε4 combinations, while those with the *APOE* ε2/ ε2, *APOE* ε2/ ε3, *APOE* ε3/ε3 were classified as non-*APOE4*-carriers. *APOE* ε2/ε4 is usually removed because it has potential risk and protective alleles[5]. We elected for an ε4 dominant (i.e., present vs. absent) rather than a dose model (i.e., 0/1/2), because there were relatively few ε4 homozygotes in our data. For analyzing the main effect of *APOE* on BAG, we further grouped *APOE* status into *APOE2* carriers (*APOE* ε2/ ε2, *APOE* ε2/ ε3), *APOE3* homozygotes (*APOE* ε3/ε3), and *APOE4* carriers (*APOE* ε3/ε4 or *APOE* ε4/ε4).

### Potential Confounders

We included potential confounding factors from baseline in UKB: continuous age, sex (females and males), education (College or University degree and others), Townsend Deprivation Index (an area-based score, continuous), household income (less than 18,000, 18,000-30,999, 31000-51,999, 52,000-99,999, more than 100,000), moderate alcohol consumption (women: ≤1 unit/day, men: ≤2 unit/day, yes/no), sedentary behavior (hours of watching television as the proxy), self-related social connection (0-active, 1-moderately active, 2/3-isolated), and baseline health conditions (yes or no: hypertension, cardiovascular diseases, type 2 diabetes, cancer, brain diseases). The missing data rate for education (6.27%), Townsend Deprivation Index (0.085%), household income (6.85%), moderate alcohol consumption (10.55%), television time (6.77%), social connection (0.36%) can be found in Table 1. To impute estimates of data that were missing at random or missing completely at random, we performed multiple imputations by chained equations (MICE)[6] to impute the missing values.


1. Wain LV, Shrine N, Miller S, et al. Novel insights into the genetics of smoking behaviour, lung function, and chronic obstructive pulmonary disease (UK BiLEVE): a genetic association study in UK Biobank. *The Lancet Respiratory Medicine.* 2015;3(10):769-781.
2. Bycroft C, Freeman C, Petkova D, et al. The UK Biobank resource with deep phenotyping and genomic data. *Nature.* 2018;562(7726):203-209.
3. Purcell S, Neale B, Todd-Brown K, et al. PLINK: a tool set for whole-genome association and population-based linkage analyses. *Am J Hum Genet.* 2007;81(3):559-575.
4. Lyall DM, Ward J, Ritchie SJ, et al. Alzheimer disease genetic risk factor APOE e4 and cognitive abilities in 111,739 UK Biobank participants. *Age Ageing.* 2016;45(4):511-517.
5. Wisdom NM, Callahan JL, Hawkins KA. The effects of apolipoprotein E on non-impaired cognitive functioning: a meta-analysis. *Neurobiol Aging.* 2011;32(1):63-74.
6. Azur MJ, Stuart EA, Frangakis C, Leaf PJ. Multiple imputation by chained equations: what is it and how does it work? *Int J Methods Psychiatr Res.* 2011;20(1):40-49.
